# Social Impacts of Multi-City HIV Research Participation Among Sexual and Gender Expansive Individuals in Kazakhstan

**DOI:** 10.1101/2024.04.22.24306169

**Authors:** Vitaliy Vinogradov, Yong Gun Lee, Gulnara Zhakupova, Gaukhar Mergenova, Alissa Davis, Emily Allen Paine, Kelsey G. Reeder, Caitlin I. Laughney, Jimin Sung, Sholpan Primbetova, Assel Terlikbayeva, Jeremy Sugarman, Elwin Wu

## Abstract

**Background:** Sexual and gender expansive (SGE) individuals in Kazakhstan are disproportionately affected by HIV yet stigma and discrimination pose ethical and practical challenges for HIV prevention research involving them. Although researchers are tasked with ensuring that risks of research participation are reasonable in relation to its benefits, participation-related risks and benefits—including negative and positive social impacts (NSIs and PSIs respectively) on personal relationships, social status, health, and other aspects of life—among SGE populations have received little attention.

**Methods:** We examined NSIs and PSIs of participation among SGE individuals in a three-city HIV prevention study in Kazakhstan at the clinical trial’s follow-up visits. We analyzed responses from 579 unique SGE participants who completed a total of 2648 follow-up visits over the 36-month study period (2019–2022).

**Results:** Overall, NSIs were rare: 9 (2%) participants reported NSIs during the study; virtually no NSIs (*x̅*=0.0037, *SD*=0.03) were reported at each follow-up visit. These few NSIs included ‘trouble with friends, family, or acquaintances’ and ‘other’. By contrast, PSIs were extensive: 515 (89%) participants reported PSIs during the study; almost an average of five PSIs (*x̅*=4.8, *SD*=3.4) were reported at each follow-up visit. The most endorsed PSIs were ‘gained knowledge’, ‘improvement in HIV-related issues’, and ‘improvement in life’.

**Conclusions:** Our findings demonstrate the potential for HIV prevention research to be associated with PSIs for SGE individuals experiencing stigmatization and discrimination. Future research should address NSIs, particularly interpersonal challenges among network members, within HIV prevention research to minimize risks and burdens of participation.

**ClinicalTrials.gov:** NCT02786615

## INTRODUCTION

Globally, the number of new HIV infections has steadily declined over the past decade.^1^ However, this progress has not been achieved equally across countries and key populations, with the number of new infections increasing in some contexts.^1,2^ In Kazakhstan, where HIV incidence grew by 88% between 2010 and 2021,^3^ sexual and gender expansive (SGE) individuals at elevated risk of HIV infection, particularly gay, bisexual, and other men who have sex with men (MSM) and transgender and nonbinary persons who have sex with men (TSM), are disproportionately affected by HIV across the HIV care continuum.^4–6^

A long line of research suggests that the enduring gaps in the HIV care continuum engagement among SGE individuals may be attributed to stigma.^7–9^ In Kazakhstan, multiple forms of stigma against SGE individuals, such as sexual and gender-based victimization in community settings, bias-based harassment in health care settings, and the absence of legal protection from discriminatory practices, have been reported.^10–13^ Moreover, among a sample of MSM and TSM in Kazakhstan, HIV stigma, internalized homophobia, and sexual and gender-based victimization and discrimination were associated with disruptions to HIV testing and treatment.^14,15^ Furthermore, SGE individuals contend with multiple behavioral risks associated with HIV that urge attention.^16–18^

Persistent stigma and discrimination pose ethical and practical challenges for research needed to help develop locally effective means of decreasing HIV incidence. For instance, HIV research participation may elicit and, in turn, be deterred by presumption of infection status, unwanted attention or isolation, and other negative attitudes toward participants.^19,20^ Further, confidentiality breach, despite efforts to mitigate it, may occur during research and could reinforce social vulnerability among participants.^21^ Although researchers are tasked with ensuring that human subjects’ risks of research participation are reasonable in relation to benefits, the risk-benefit relationship may not always be evident.^22^ These risks and benefits include negative (NSIs) and positive social impacts (PSIs) pertaining to personal relationships, social status, health, and other aspects of participants’ lives.^23^ Evidence in other settings suggests that PSIs related to participation in HIV prevention research are prevalent, whereas NSIs are uncommon.^24–27^ Still, scientific literature on the assessment of NSIs and PSIs of HIV prevention research participation among SGE individuals, particularly those in high stigma contexts, is sparse. We sought to expand on this work by examining the overall incidence and correlates of NSIs and PSIs related to HIV prevention research participation among a sample of MSM and TSM in Kazakhstan.

## METHODS

We used data from a stepped wedge clinical trial of an HIV preventive intervention for MSM and TSM in Kazakhstan (NCT02786615). Briefly, the intervention involved crowdsourcing and peer-actuated network approaches to increasing engagement of MSM and TSM in the HIV care continuum in Kazakhstan.^28^ After a period of collecting baseline data at participants’ initial visits (July 2018 – February 2019), the intervention was implemented sequentially, across three Kazakhstan cities: Almaty, Shymkent, and Astana. During the implementation phase, participants were invited to complete a manualized intervention that was delivered over four sessions preceded by an orientation session.^29^

Throughout the trial, we conducted robust community engagement including community advisory board sessions and recruitment venue site visits to help ensure safety and comfort in research participation. This effort engaged community stakeholders in sharing their knowledge and wisdom of best practices in conducting research involving SGE individuals in Kazakhstan. For instance, we inquired each participant about preferred/appropriate term(s) to describe members of SGE communities and learned that the term oft-used in research, ‘sexual and gender minority’, likely carries negative connotations in local dialect. Finally, we implemented safety plans and staff training for risk mitigation, health care and social service referrals, and regularly monitored social impacts during the trial.

### Participants

Eligibility criteria for the study included: being ≥18 years old; identifying as man or being assigned male at birth; reporting consensual sex with a man in the past 12 months; reporting binge drinking, drug use, or both in the past 90 days; and residing in a study city. For the purposes of the study, we refer to participants as ‘MSM’, ‘TSM’, or ‘SGE individuals’, acknowledging the terms’ limitation in thoroughly representing sexual and gender diversity among them.

### Procedures

Participants provided informed consent at their initial study visit. A structured behavioral survey was administered by trained Research Assistants at the initial visit and follow-up visits every 6 months until the end of the trial. All survey items were developed in English, translated to Russian and Kazakh, and back-translated to English to ensure accuracy. Due to the stepped wedge study design, while most participants completed six follow-up visits, those who were enrolled earlier in the trial had more follow-up visits scheduled compared to those enrolled later/closer to the end of the trial. Rapid HIV testing, counseling, and confirmatory testing or treatment referral for HIV, syphilis, gonorrhea, and chlamydia were offered at the initial visit; the same services for HIV were offered at the final follow-up visit. Study procedures were reviewed and approved by the Institutional Review Boards at Columbia University and Al-Farabi Kazakh National University.

Between July 2018 and February 2022, 982 individuals were screened; of them, 649 (66%) were determined eligible for participation in the study. Of the study-eligible individuals, 629 (97%) returned for the initial visit; of them, 579 (92%) were subsequently retained for at least one follow-up visit. Social impacts related to HIV prevention research participation were assessed at follow-up visits. For this study, we used panel data from 579 participants who completed a total of 2,648 follow-up visits over a 36-month period (February 2019 – February 2022).

### Measures

#### Social Impact

Negative social impact (NSI) and positive social impact (PSI) items were adapted from the social impact assessment questionnaire used in a multinational HIV prevention study involving people who inject drugs.^30^ For NSIs, participants were asked at each follow-up visit whether anything negative or risky happened to them due to their study participation. Those reporting any negative incident were probed about occurrences of specific NSIs (1=yes, 0=no). Similarly, for PSIs, participants were asked whether anything positive or beneficial happened due to their study participation. Those reporting any positive incident were probed about occurrences of specific PSIs (1=yes, 0=no).

#### Alcohol and Drug Use

Alcohol and drug use assessment relied on the same measures used in previous studies.^14,16,17^ Briefly, participants self-reported lifetime use of alcohol, marijuana, heroin and other opioids, stimulants, cocaine, hallucinogens or psychedelics, inhalants, and club drugs. Participants reporting any prior use of alcohol were asked about incidents of binge drinking, operationalized as “consuming five or more alcohol beverages within a two-hour period”.^31^ When appropriate, local or street names were used to describe the drugs. For alcohol, binge drinking, and each of the drugs, participants reporting any prior use were probed about the frequency of use in the past 90 days.

#### Sociodemographic Characteristics

Participants self-reported their city of residence, age (in years), legal marital status, education completion status, employment status, gender identity, sex assigned at birth, sexual orientation, and HIV status. Participants reporting ‘other’ were asked to specify their responses.

### Study Variables and Data Analysis

Social impacts were analyzed in two ways (1) *per visit*, in which each follow-up visit constitutes a case, and (2) *per participant*, in which each participant’s series of follow-ups constitutes a case. In other words, the outcome data were structured in long format and wide format respectively, with long format allowing for easy calculation of metrics using visits as the unit of analysis, and wide format involves using study participants as the unit of analysis. For each set of NSI and PSI items in the long format data, responses were summed to compose count variables of NSI and PSI items endorsed at each follow-up visit; these variables were dichotomized to create dummy variables denoting any or no social impact. Next, we transformed long format data into wide format. For each set of NSI and PSI items in the wide format data, we calculated the averages of count variables across completed follow-up visits; these average count variables were dichotomized to create dummy variables so that the value of zero denoted no social impact, while values greater than zero denoted any social impact. Finally, for each social impact item, responses across completed follow-up visits were summed and dichotomized to create a dummy variable denoting any or no social impact during study participation.

Analysis of alcohol and drug use focused on use that was inconsistent with medical or legal guidance, and thus excluded alcohol use and included binge drinking and drug use. Responses for binge drinking were imputed as zero if no prior alcohol use was reported. Frequencies of binge drinking and drug use in the past 90 days were dichotomized to create dummy variables denoting any or no recent use. Frequencies were imputed as zero if no prior use was reported. The dichotomized responses for drug use across all drugs were summed and dichotomized to create a dummy variable denoting any or no drug use.

Responses for age were dichotomized as ‘ages 18-24’ and ‘ages ≥25’ categories based on clinical and cultural relevance.^32,33^ If feasible and appropriate, write-in responses for the ‘other’ query for the sociodemographic characteristic items were inductively coded as existing response options, as determined by the authors (VV, YL, EW). In this way, all descriptions for the ‘other’ sexual orientation category were coded as ‘bisexual.’ At the initial visit, no participant reported being assigned ‘other’ sex at birth.

We conducted frequency analyses to describe the distributions of sociodemographic characteristics and alcohol and drug use at the initial visit, as well as NSI and PSIs at follow-up visits. Additionally, we performed linear regression analyses to test bivariate associations between sociodemographic and alcohol and drug use factors at the initial visit and the average count of PSIs endorsed across follow-up visits. We did not perform bivariate analyses for NSIs due to very small numbers of NSIs reported (as presented in Results). All statistical analyses were performed using SPSS version 28.0 (IBM Corp., Armonk, NY).

## RESULTS

Descriptive statistics for the initial-visit sociodemographic characteristics and alcohol and drug use of the study sample of MSM and TSM who provided follow-up data are presented in Table 1. Participants were between the ages of 18 and 59 (median=27). Most participants were single (79%), employed (77%), and had completed vocational or college/university-level education (80%). With respect to sexual and gender identities, most identified as man (90%) and sexually expansive, i.e., gay (53%) or bisexual (46%). In this sample, 489 (85%) and 252 (44%) reported recent binge drinking and drug use, respectively. A quarter of the sample reported unknown HIV status.

**Table 1.** Sociodemographic and Substance Use Characteristics a Sample of MSM and TSM in Kazakhstan at Initial Visit of an HIV Prevention Clinical Trial (*N*=579)

| Variable | n (%) |
| --- | --- |
| City of Residence |  |
| Almaty | 227 (39.2%) |
| Shymkent | 184 (31.8%) |
| Astana | 168 (29.0%) |
| Age groups, years (median=27; range=18-59) |  |
| 18-24 | 207 (35.8%) |
| 25 or older | 372 (64.2%) |
| Legal Marital Status |  |
| Single, never married | 455 (78.6%) |
| Else (e.g., married, no longer with spouse, other) | 124 (21.4%) |
| Education Completion Status |  |
| High school | 116 (20.0%) |
| Vocational or college/university | 463 (80.0%) |
| Employment Status |  |
| Employed (full-time or part-time) | 447 (77.2%) |
| Unemployed | 63 (10.9%) |
| Else (e.g., retired, disability, homemaker, student, other) | 69 (11.9%) |
| Gender Identity |  |
| Woman | 32 (5.5%) |
| Man | 521 (90.0%) |
| Other | 26 (4.5%) |
| Sex Assigned at Birth |  |
| Female | 2 (0.3%) |
| Male | 577 (99.7%) |
| Sexual Orientation |  |
| Straight | 10 (1.7%) |
| Gay | 305 (52.7%) |
| Bisexual | 264 (45.6%) |
| Self-Reported HIV Status |  |
| Positive | 46 (7.9%) |
| Negative | 387 (66.8%) |
| Unknown | 146 (25.2%) |
| Drug Use, Lifetime | 432 (74.6%) |
| Drug Use, Past 90 Days | 252 (43.5%) |
| Binge Drinking, Lifetime | 527 (91.0%) |
| Binge Drinking, Past 90 Days | 489 (84.5%) |

**Table 2.** Negative Social Impacts and Positive Social Impacts of HIV Research Participation in a Sample of MSM and TSM in Kazakhstan (*N*=2648 visits)

| **2a. Negative Social Impacts (NSIs)** |  |
| --- | --- |
| **Variable** | ***n* (%)** |
| Any NSI | 9 (0.3%) |
| Items |  |
| Other NSI | 6 (0.2%) |
| Trouble with friends, family, or acquaintances | 5 (0.2%) |
| Arrested or trouble with police or other legal problems | 0 |
| Trouble getting or keeping housing | 0 |
| Trouble getting or keeping job, income, or economic support | 0 |
| Trouble getting health care or health insurance | 0 |
| **2b. Positive Social Impacts (PSIs)** |  |
| **Variable** | ***n* (%)** |
| Any PSI | 1519 (57.4%) |
| Items |  |
| Gained knowledge | 1445 (54.6%) |
| Improvement in HIV-related issues | 1428 (53.9%) |
| Improvement in mental health | 1162 (43.9%) |
| Improvement in life | 1151 (43.5%) |
| Improvement in physical health | 1133 (42.8%) |
| Improvement in (connection to) community | 1107 (41.8%) |
| Improvement in relationships | 1060 (40.0%) |
| Less homophobia or improvement in handling homophobia better | 940 (35.5%) |
| Reduction of stigma | 894 (33.8%) |
| Improvement in financial | 770 (29.1%) |
| Improvement in employment | 667 (25.2%) |
| Reduction in drug use | 491 (18.5%) |
| Other PSI | 457 (17.3%) |
| Less transphobia or improvement in handling transphobia better | 408 (15.4%) |

**Table 3.**
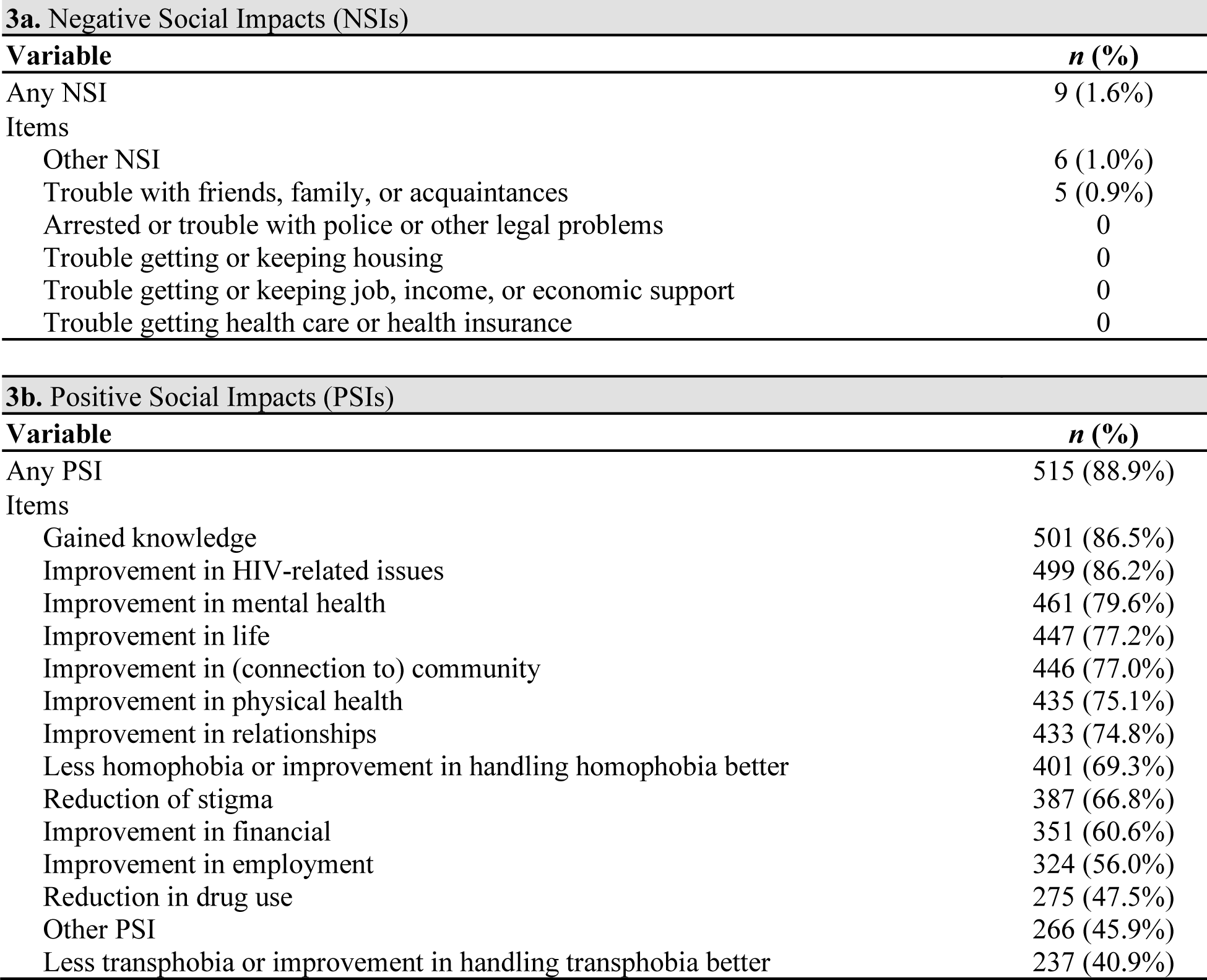
Negative Social Impacts and Positive Social Impacts of HIV Research Participation in a Sample of MSM and TSM in Kazakhstan (*N*=579 participants)

A graphical summary of participant responses for NSIs and PSIs is presented in Figure 1. In this raster format, each row (i.e., raster line) represents a participant and the raster line is divided into columns, with each column indicating whether NSIs and PSIs (Figures 1a and 1b, respectively) were reported for the corresponding follow-up visit. The greater the number of NSIs (out of 6 items) or PSIs (out of 14 items) reported during each visit, the more saturated the color (red for NSIs in Figure 1a and green for PSIs in Figure 1b). If a participant did not complete a particular visit due to missing a scheduled appointment, the follow-up visit falling after the end of the trial, or any other reason, that is indicated with a yellow element in the raster line. Overall, the plots depict a higher volume of more-saturated green raster lines than saturated red lines, indicating many more PSIs than NSIs were reported by participants at each follow-up visit.

**Figure 1.**
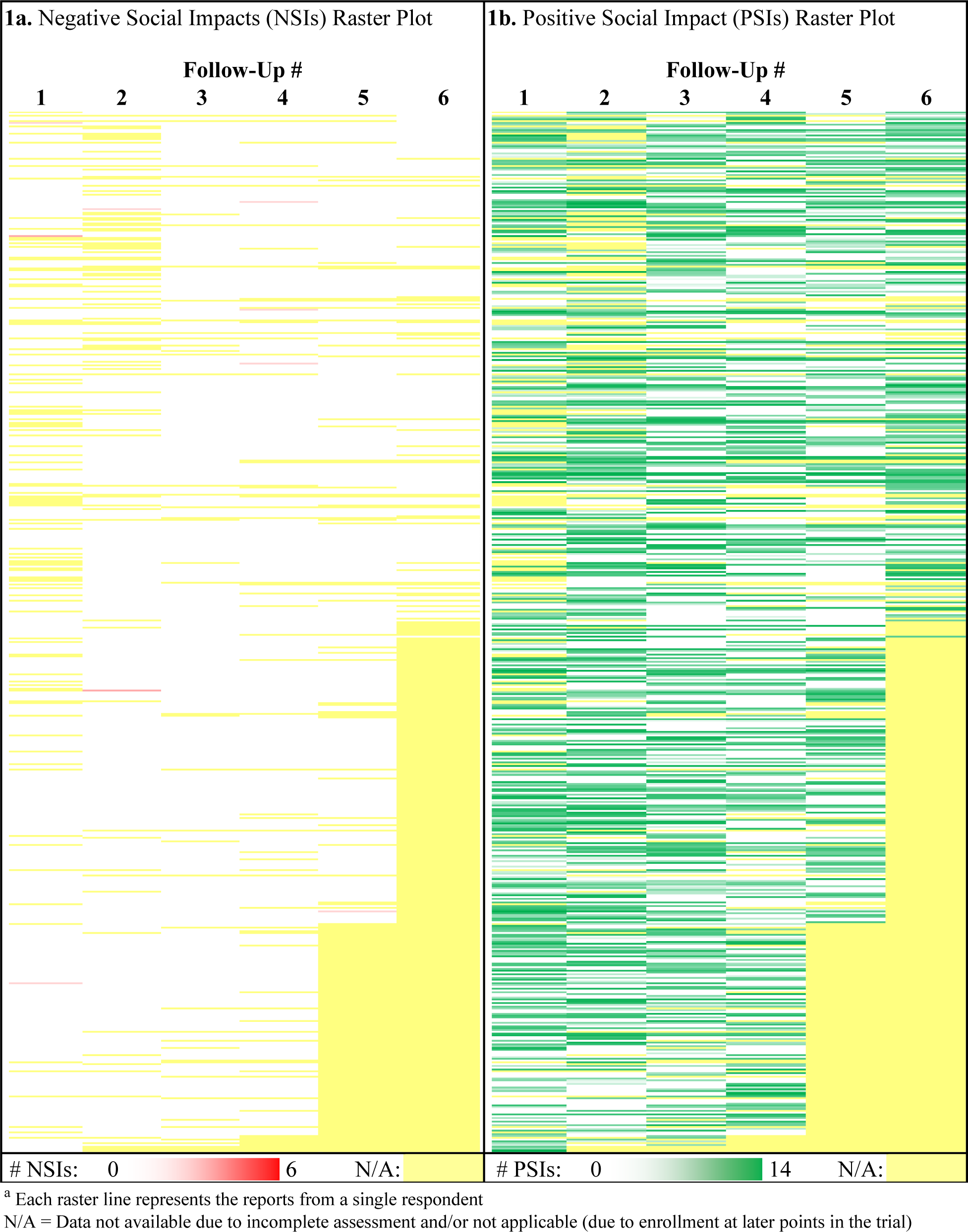
Raster^a^ Plots of Negative Social Impacts and Positive Social Impacts of HIV Research Participation in a Sample of MSM and TSM in Kazakhstan (*N*=2648)

### Social Impacts Per Visit

Metrics regarding the distributions of NSIs and PSIs using the long format are presented in Table Of 2648 follow-up visits, nine (0.3%) had a report of at least one NSI. Specifically, there were five (0.2%) reports of ‘trouble with friends, family, or acquaintances’ and six (0.2%) reports of ‘other NSI’ involving ‘loss of privacy’ (*n*=4), ‘stigma/harassment due to study participation’ (*n*=1), and ‘trouble with HIV-related issues’ (*n*=1). By contrast, 1519 visits (57%) had respondents reporting at least one PSI.

### Social Impacts Per Participant

Metrics regarding the distributions of NSIs and PSIs using the wide format are presented in Table Of 579 MSM and TSM, nine individuals (2%) reported an NSI at least once across follow-up visits; five (1%) reported ‘trouble with friends, family, or acquaintances’ and six (1%) reported ‘other NSI’. There was no report of NSIs related to legal, housing, economic, or health care issues. By contrast, 515 individuals (89%) reported a PSI at least once across follow-up visits. The majority of participants reported ‘gained knowledge’ (87%), ‘improvement in HIV-related issues’ (86%), and ‘improvement in life’ (80%); fewer participants reported ‘less transphobia or improvement in handling it better’ (41%), ‘reduction in drug use’ (48%), and ‘other PSI’ (46%), but these numbers still approached approximately half of participants. Participants reported, on average, five PSIs (*x̅*=4.8, *SD*=3.4) and zero NSI (*x̅*=0.0037, *SD*=0.03) during the trial.

### Correlates of Positive Social Impacts

Partial results of the bivariate analyses illustrating statistically significant correlates of the average count of PSIs—residence and recent binge drinking—are presented in Table 4. On average, living in Shymkent and Astana, compared with Almaty, was associated with 2.8-point and 1.3-point higher average count of PSIs, respectively. Recent binge drinking was associated with, on average, a 1.2-point higher average count of PSIs.

**Table 4.** Significant Bivariate Associations Between the Average Count of Positive Social Impacts Endorsed Across Follow-Up Visits and Sociodemographic and Substance Use Characteristics at Initial Visit in a Sample of MSM and TSM in Kazakhstan (*N*=579)

| Variable | $\beta$ | $SE$ | 95% $CI$ | $p$ |
| --- | --- | --- | --- | --- |
| City of Residence |  |  |  |  |
| Almaty | Ref. |  |  |  |
| Shymkent | 2.8 | 0.3 | (2.2, 3.5) | <.001 |
| Astana | 1.3 | 0.3 | (0.7, 1.9) | <.001 |
| Binge Drinking, Past 90 Days | 1.2 | 0.4 | (0.4, 2.0) | .002 |

*SE*: Standard Error
*CI*: Confidence Interval

## DISCUSSION

Among MSM and TSM enrolled in an HIV prevention clinical trial in Kazakhstan, PSIs related to study participation were endorsed at substantially higher rates than NSIs. Specifically, NSIs were very infrequently reported, with an average of virtually zero NSI per follow-up visit and per participant, whereas PSIs were reported extensively, with an average of five PSIs per visit. To our knowledge, this is the first study to examine the social impacts of HIV prevention research participation among SGE individuals in Kazakhstan.

In our study, the most frequently endorsed PSIs were ‘gained knowledge’, ‘improvement in HIV-related issues’, and ‘improvement in mental health’. The parent study was testing the efficacy of a social network-based intervention for increasing engagement of MSM and TSM in the HIV care continuum in Kazakhstan. A primary objective of the intervention was supporting MSM and TSM with promoting the uptake of HIV care services within their networks. Therefore, receipt of or indirect exposure to the intervention could have contributed to improvement of knowledge and HIV-related issues among participants.

Our findings are consistent with other studies demonstrating common experiences of PSIs and scant reports of NSIs. A multinational HIV prevention trial involving people who inject drugs in China and Thailand similarly found that 77% of participants reported PSIs, far exceeding 0.5% reporting NSIs.^24^ A cross-protocol analysis of data from 413 preventive HIV vaccine trials in 13 countries showed that 81% of participants reported PSIs and 8% reported NSIs.^25^ Finally, an HIV cure-related trial involving women living with HIV in the U.S. demonstrated reporting of a greater number of PSIs than NSIs by participants at their exit survey.^26^ Our study extends this line of inquiry by contributing analyses of responses from SGE individuals in Kazakhstan.

In our study, participants reporting recent binge drinking were more likely to report a greater number of PSIs. Binge drinking is associated with increased risk of developing health and psychosocial problems. Since the trial intervention also focused on increasing engagement in alcohol and drug use treatment, it is possible that participants reporting binge drinking perceived the intervention to be beneficial and valuable more so than those reporting no binge drinking. Moreover, participants living in Shymkent and Astana were more likely to report a greater number of PSIs. In previous studies, geographical differences in PSI endorsement had been attributed to variability in access to resources.^25,30^ Given that twice as many MSM and TSM were estimated to be living in Almaty as Shymkent or Astana, it is reasonable to posit that participants in Almaty may have been benefiting from having greater availability of and access to resources outside the study.^5^

Although relatively rare, NSIs warrant careful review and, as necessary, development of mitigation strategies. In our study, the most frequently endorsed NSIs were ‘other NSI’ and ‘trouble with friends, family, or acquaintances’. Other NSIs mostly entailed loss of privacy due to breach of confidentiality by an entity (e.g., HIV care provider) or an individual—in at least one case, by another study participant. One participant in our study recounted online criticisms about their study participation. Breach of confidentiality had been reported as an NSI in a multinational preventive HIV vaccine trial.^25^ In addition, navigating unwanted attention or aggression regarding study participation through confrontation or avoidance had been reported in other studies,^22,26^ and could be interpreted as trouble with personal relationships. Indeed, in our study, trouble with friends, family, or acquaintances was endorsed by five out of nine participants reporting any NSI. Although we confirmed with participants that NSIs had not resulted due to administration of study procedures, they nonetheless stress the importance of developing and implementing safety plans to mitigate potential NSIs.

Limitations to our study can inform future research. In this study, assessment of NSIs and PSIs pertained to experiences of research study participation and did not specify a timeframe. Responses at subsequent follow-up visits could reflect prior experiences and, therefore, lead us to capture individual PSIs and NSIs multiple times. This limitation does not impact participant-level results; however, future studies interested in time-sequencing of NSIs and PSIs can overcome this limitation by specifying a timeframe, e.g., since the most recent prior study visit. Also, given that our social impact questionnaires reflected categories of impacts that could be interpreted in different ways, future research should cognitively test the questionnaire items (following translation) to ensure accuracy in the local context. Next, endorsement of social impacts may be subject to social response bias. In other settings, it has been speculated that fear of losing tangible benefits, such as medical care and compensation, could lead to underreporting of NSIs.^25^ It is possible that study participants underreported NSIs to avoid losing HIV/STI testing and counseling/referral, compensation provided for their participation, or both. Training of staff on data collection procedures ensuring response accuracy should be prioritized. Finally, our findings may not be generalizable to all MSM and TSM in Kazakhstan. This analysis was restricted to data provided by SGE individuals retained in an HIV prevention trial conducted in three Kazakhstan cities over a three-year period and, thus, did not represent experiences of those who did not participate in or had been excluded from the trial (e.g., those without recent drug use or binge drinking). However, we recruited a large sample of MSM and TSM from multiple cities in Kazakhstan (the first study to do so to our knowledge) and retention rates were high, enabling us to obtain diverse perspectives on NSI and PSI from this population. Moreover, given that drug use and binge drinking are also stigmatized, participants in our study likely experience more stigma compared to their peers who do not engage in these behaviors—lending confidence to our conclusion that HIV prevention research overwhelmingly leads to PSIs in a high-stigma context. At the same time, despite extensive retention efforts, missing data were incurred through participant dropout and loss to follow-up. It is possible that attrition was related to anticipation of NSI or absence of PSI. As such, future studies specifically designed to assess PSIs and NSIs among MSM and TSM in high-stigma contexts is warranted to bolster assessment of ethics of conducting HIV prevention research among stigmatized groups.

## CONCLUSIONS

Our study adds to the literature on social impact assessment in HIV prevention trials and highlights the ethical significance of assessing participant-centered experiences. PSIs and NSIs identified in this study suggest that it is possible to ethically conduct HIV prevention research involving SGE individuals at elevated risk of HIV infection in Kazakhstan despite the context of intersecting stigma. As risk mitigation is an important ethical consideration in HIV prevention, the infrequent reporting of NSIs in our study is encouraging. It is possible that procedures embedded in our trial to anticipate and address ethical issues contributed to such an outcome. These procedures included, but were not limited to, involving SGE individuals, as paid staff, or community advisory board members, in all aspects of the trial, training staff on protecting the safety, privacy, and confidentiality of participants, creating an affirming environment for research participation, hosting social events for building rapport and comfort, and reviewing and sharing study findings with SGE communities and service providers.

Several recommendations are derived from this study: (1) partner with SGE communities in all aspects of research; (2) develop safety plans, including risk mitigation strategies, tailored to the needs of SGE communities and features of research settings/platforms;^34,35^ (3) embed systematic assessment of social impacts related to study participation; (4) include PSIs in evaluating the ethics of conducting research;^36^ and (5) facilitate feedback and communication with SGE communities to ensure researcher accountability. These recommendations may serve as a guide for conducting HIV prevention research in settings where diverse sexual and gender characteristics are stigmatized with few NSIs and many PSIs.

## STATEMENTS AND DECLARATIONS

## Acknowledgements

We would like to thank the project staff of the Columbia University Global Health Research Center of Central Asia—including Karina Alipova, Farruh Aripov, Olga Balabekova, Daniya Bekishev, Dilara Belkesheva, Valeriya Davydova, Ferangiz Hasanova, Sultana Kali, Saltanat Kuskulova, Aitkul Nazarova, Syrym Omirbek, Svyatoslov Suslov, Aizhan Toleuova, Aidar Yelkeyev, and Saida Yessenova—for their commitment and care given to the execution of study activities. We would like to thank the participants for their courage and time in sharing their stories.

## Data availability

Deidentified data used in this study could be made available from the corresponding author upon reasonable request.

## REFERENCES

1. Joint United Nations Programme on HIV/AIDS (UNAIDS). UNAIDS Data 2023. 2023. Available at: https://www.unaids.org/en/resources/documents/2023/2023_unaids_data.

2. Joint United Nations Programme on HIV/AIDS (UNAIDS). Miles to go - The response to HIV in eastern Europe and central Asia. 2018. Available at: https://www.unaids.org/en/resources/documents/2018/miles-to-go_eastern-europe-and-central-asia.

3. Joint United Nations Programme on HIV/AIDS (UNAIDS). UNAIDS Data 2022. 2022. Available at: https://www.unaids.org/en/resources/documents/2023/2022_unaids_data.

4. Eurasian Coalition on Male Health (ECOM). Brief on HIV among MSM in Kazakhstan. 2018. Available at: https://ecom.ngo/library/brief-kazakhstan-en.

5. Wu E, Terlikbayeva A, Hunt T, Primbetova S, Lee YG, Berry M. Preliminary population size estimation of men who have sex with men in Kazakhstan: Implications for HIV testing and surveillance. LGBT Health. 2016;4:164–167. doi:10.1089/lgbt.2015.0152

6. Wu E, Lee YG, Zhakupova G, et al. Prevalence and correlates of HIV infection and unknown HIV infection among men who have sex with men (MSM) in Kazakhstan: Evidence for a brewing syndemic from a multi-city study. Presented at: 23rd International AIDS Conference (AIDS 2020); 2020; Virtual.

7. Arreola S, Santos G-M, Beck J, et al. Sexual stigma, criminalization, investment, and access to HIV services among men who have sex with men worldwide. AIDS Behav. 2015;19:227–234. doi:10.1007/s10461-014-0869-x

8. Babel RA, Wang P, Alessi EJ, Raymond HF, Wei C. Stigma, HIV risk, and access to HIV prevention and treatment services among men who have sex with men (MSM) in the United States: A scoping review. AIDS Behav. 2021;25:3574–3604. doi:10.1007/s10461-021-03262-4

9. Vaitses Fontanari AM, Zanella GI, Feijó M, Churchill S, Rodrigues Lobato MI, Costa AB. HIV-related care for transgender people: A systematic review of studies from around the world. Soc Sci Med. 2019;230:280–294. doi:10.1016/j.socscimed.2019.03.016

10. ALMA-TQ, Law CfIHRoNPSo, Rights GIfH. Violations by Kazakhstan of the Right of Transgender Persons to Legal Recognition of Gender Identity. 2016. Available at: https://www.kok.team/files/files/article/386/int-ccpr-css-kaz-24305-e.pdf

11. Eurasian Coalition on Male Health (ECOM). Legislative Analaysis Related to SOGI and HIV in Kazakhstan. 2021. Available at: https://ecom.ngo/library/kazakhstan-legislative-analysis_2020_en

12. Levitanus M. Agency and resistance amongst queer people in Kazakhstan. Centr Asian Surv. 2022;41:498–515. doi:10.1080/02634937.2021.2008874

13. Laughney CI, Lee YG, Mergenova G, et al. Earlier sexual debut and anti-gay victimization among men who have sex with men (MSM) in Kazakhstan. J Interpers Violence. 2023;38:10795–10813. doi:10.1177/08862605231176800

14. Paine EA, Lee YG, Vinogradov V, et al. HIV Stigma, homophobia, sexual and gender minority community connectedness and HIV testing among gay, bisexual, and other men and transgender people who have sex with men in Kazakhstan. AIDS Behav. 2021;25:2568–2577. doi:10.1007/s10461-021-03217-9

15. Paine EA, Lee YG, Mergenova G, et al. Compounding vulnerabilities: Victimization and discrimination is associated with COVID-19 disruptions to HIV-related care among gay, bisexual, and other men and transgender and nonbinary people who have sex with men in Kazakhstan. AIDS Care. 2023;35:651–657. doi:10.1080/09540121.2022.2148956

16. Lee YG, Zhakupova G, Vinogradov V, et al. Polydrug use, sexual risk, and HIV testing among cisgender gay, bisexual, and other men and transgender and nonbinary individuals who have sex with men in Kazakhstan. AIDS Educ Prev. 2022;34:413–426. doi:10.1521/aeap.2022.34.5.413

17. Laughney CI, Lee YG, Mergenova G, et al. Earlier sexual debut as a risk factor for substance use among men who have sex with men (MSM) in Kazakhstan. Glob Soc Welf. 2023;doi:10.1007/s40609-023-00298-3

18. Laughney CI, Lee YG, Mergenova G, et al. Earlier sexual debut and exchange sex among men who have sex with men (MSM) in Kazakhstan. J Sex Res. 2023;60:919–924. doi:10.1080/00224499.2023.2167064

19. MacQueen K, Shapiro K, Karim Q, Sugarman J. Ethical challenges in international HIV prevention research. Account Res. 2004;11:49–61. doi:10.1080/08989620490280230

20. Bass SB, D’Avanzo P, Alhajji M, et al. Exploring the engagement of racial and ethnic minorities in HIV treatment and vaccine clinical trials: A scoping review of literature and implications for future research. AIDS Patient Care STDs. 2020;34:399–416. doi:10.1089/apc.2020.0008

21. Azhar S, Tao X, Jokhakar V, Fisher CB. Barriers and facilitators to participation in Long-acting injectable PrEP research trials for MSM, transgender women, and gender-nonconforming people of color. AIDS Educ Prev. 2021;33:465–482. doi:10.1521/aeap.2021.33.6.465

22. Gilbertson A, Kelly EP, Rennie S, Henderson G, Kuruc J, Tucker JD. Indirect benefits in HIV cure clinical research: A qualitative analysis. AIDS Res Hum Retroviruses. 2018;35:100–107. doi:10.1089/aid.2017.0224

23. Allen M, Lau C-Y. Social impact of preventive HIV vaccine clinical trial participation: A model of prevention, assessment and intervention. Soc Sci Med. 2008;66:945–951. doi:10.1016/j.socscimed.2007.10.019

24. Sugarman J, Stalter R, Bokoch K, Liu T-Y, Donnell D. Positive social impacts related to participation in an HIV prevention trial involving people who inject drugs. IRB. 2015;37:17–19.

25. Andrasik MP, Sesay FA, Isaacs A, Oseso L, Allen M. Social impacts among participants in HIV Vaccine Trial Network (HVTN) preventive HIV vaccine trials. J Acquir Immune Defic Syndr. 2020;84:488–496. doi: 10.1097/QAI.0000000000002369

26. Dubé K, Hosey L, Starr K, et al. Participant perspectives in an HIV cure-related trial conducted exclusively in women in the United States: Results from AIDS clinical trials group 5366. AIDS Res Hum Retroviruses. 2020;36:268–282. doi:10.1089/aid.2019.0284

27. Dubé K, Shelton B, Patel H, et al. Perceived risks and benefits of enrolling people with HIV at the end of life in cure research in Southern California, United States. J Virus Erad. 2023;9:100328. 10.1016/j.jve.2023.100328

28. Wu E. Using social media and social networks to increase access to HIV care among MSM in three cities in Kazakhstan: Project UNI. Presented at: 22nd International AIDS Conference (AIDS 2018); 2018; Amsterdam, Netherlands.

29. Wu E, Lee YG, Vinogradov V, et al. Intervention adaptation and implementation method for real-world constraints and using new technologies. Res Soc Work Pract. 2022;33:562–570. doi:10.1177/10497315221120605

30. Sugarman J, Trumble I, Hamilton E, et al. Reported participation benefits in international HIV prevention research with people who inject drugs. Ethics Hum Res. 2019;41:28–34. doi:10.1002/eahr.500030

31. National Institute on Alcohol Abuse and Alcoholism. Drinking Levels Defined. Available at: https://www.niaaa.nih.gov/alcohol-health/overview-alcohol-consumption/moderate-binge-drinking

32. Berry M, Wirtz AL, Janayeva A, et al. Risk factors for HIV and unprotected anal intercourse among men who have sex with men (MSM) in Almaty, Kazakhstan. PLoS One. 2012;7:e43071. doi:10.1371/journal.pone.0043071

33. World Health Organization (WHO). Adolescent and Young Adult Health. Available at: https://www.who.int/news-room/fact-sheets/detail/adolescents-health-risks-and-solutions

34. Fisher CB, Bragard E, Bloom R. Ethical considerations in HIV eHealth intervention research: Implications for informational risk in recruitment, data maintenance, and consent procedures. Curr HIV/AIDS Rep. 2020;17:180–189. doi:10.1007/s11904-020-00489-z

35. Sugarman J, Barnes M, Rose S, et al. Development and implementation of participant safety plans for international research with stigmatised populations. Lancet HIV. 2018;5:e468–e472. doi:10.1016/S2352-3018(18)30073-0

36. Rennie S, Day S, Mathews A, et al. The role of inclusion benefits in ethics committee assessment of research studies. Ethics Hum Res. 2019;41:13–22. doi:10.1002/eahr.500015

